# Evaluation of an eHealth Assistive Device Provision Training Tool: A Quasi-Experimental Study

**DOI:** 10.64898/2026.09.20.26363517

**Authors:** Rachel Sudhakar, Katrine Sauvé-Schenk, Jennifer O’Neil

**Affiliations:** Faculty of Science, University of Ottawa, Ottawa, Ontario, Canada; School of Rehabilitation Sciences, Faculty of Health Sciences, University of Ottawa, Ottawa, Ontario, Canada

## Abstract

**Background:** Approximately 2.5 billion people require assistive products (APs), yet access remains limited partly because of shortages of trained personnel. eHealth tools may help strengthen workforce capacity and translate evidence into practice. The World Health Organization’s Training in Assistive Products (TAP) program is an open-access eHealth tool designed to strengthen AP provision. However, its impact on learner outcomes remains insufficiently evaluated. This study assessed the usability and acceptability of the TAP module and changes in self-efficacy in mobility aid provision.

**Methods:** We conducted a quasi-experimental pre-post study with health science students who completed the WHO TAP “Introduction to Assistive Products” module. We assessed usability using the System Usability Scale, acceptability using the Technology Acceptance Model and Theoretical Framework of Acceptability, and self-efficacy using an adapted version of Bandura’s Multidimensional Scales of Perceived Self-Efficacy. We analyzed data using descriptive statistics and paired-samples t-tests.

**Results:** Of 53 participants, 33 completed both assessments. Usability exceeded established benchmarks (76.97 ± 9.66), indicating a positive user experience. Acceptability findings demonstrated favourable responses across domains, with high ratings for ease of use and intervention coherence, and comparatively lower burden scores. Mean self-efficacy increased across all competencies from 2.32 ± 0.58 before the module to 4.23 ± 0.36 afterward (p < 0.001), reflecting substantial gains in confidence in AP provision.

**Conclusion:** The eHealth TAP module demonstrated above-average usability and favourable acceptability, while significantly improving self-efficacy in AP provision. These findings highlight the potential for eHealth training implementation to strengthen workforce capacity and bridge training gaps in global health.

## Background

The World Health Organization (WHO) estimates that over two billion people worldwide require at least one assistive product (AP), yet only one in ten has access to them.^1^ Mobility APs, including wheelchairs, canes, and walkers, are tools that support independence, safety, and social participation for individuals with physical limitations.^2^ Proper mobility aid provision requires assessing user needs, selecting contextually appropriate APs, ensuring proper fit, and providing follow-up to monitor outcomes.^1^ Access to personnel trained to appropriately select, fit, and provide mobility aids remains limited, particularly in low- and middle-income countries, partly because of rehabilitation workforce shortages and limited standardized training.^3–5^ Inappropriate provision may result in discomfort, injury, or device abandonment.^6^ Although TAP was developed to address training gaps, individual TAP modules have received limited evaluation of learner-centred outcomes, including usability, acceptability, and self-efficacy.^7^

### eHealth TAP Training Modules

eHealth modules, defined as digitally and remotely delivered, internet-based learning tools that provide structured health-related education and training, offer opportunities for learners to gain clinical knowledge and practical understanding without the constraints of time, travel, or classroom space.^8^ Studies have shown that eHealth and hybrid learning approaches can be as effective as traditional in-person instruction when designed with learner engagement in mind.^9^ eHealth delivery enables flexible engagement and opportunities for reinforcement, which may support skill development and knowledge retention.^10^ The WHO TAP program is a free, multilingual eHealth open-access platform designed to train community and primary-level health personnel in the safe and effective delivery of APs.^11^ The modules are interactive and self-paced, combining text, visuals, and quizzes that walk learners through the four-step process of Select, Fit, Use, and Follow-up. TAP has strong potential to support workforce training, particularly for learners with limited access to hands-on rehabilitation education.

### Self-efficacy, Usability, and Acceptability in the Provision of APs

Self-efficacy refers to an individual’s belief in their ability to plan and carry out the actions needed to achieve specific goals.^12^ In health science education, self-efficacy is closely linked to skill development, learning motivation, and readiness for clinical use as perceptions of capability can influence the degree to which individuals engage with and persist in new learning tasks.^13^ Stronger self-efficacy is associated with greater confidence with complex patient scenarios, adaptability to new challenges, and persistence in skill acquisition.^14^ Evaluating self-efficacy following exposure to the eHealth TAP training module provides insight into learners’ perceived preparedness to translate knowledge into practice.

Usability and acceptability are critical factors influencing how people engage with digital learning tools. Usability refers to the extent to which a digital system enables users to achieve their goals effectively, efficiently, and with satisfaction and minimal cognitive effort.^15^ A highly usable eHealth module ensures that learners can navigate content intuitively and remain focused on the learning objectives rather than technical barriers. Acceptability refers to the extent to which an intervention is perceived by its intended users as appropriate, engaging, and satisfactory and is shaped by both cognitive and emotional responses.^16^ In eHealth education, acceptability reflects how learners perceive the value, relevance, and overall experience of a training tool, including whether they find it useful, easy to use, and worth engaging with.^17^ Therefore, we evaluated the usability and acceptability of the WHO TAP module and examined changes in participants’ self-efficacy regarding mobility aid provision.

## Methods

We followed the Knowledge-to-Action (KTA) framework, which outlines a structured process for evaluating the implementation of innovation into practice.^18,19^ Specifically, this study aligns with the monitoring and evaluating steps of the action cycle, evaluating the WHO TAP module within a controlled, university-based setting. Ethics approval for this study was obtained from the University of Ottawa Research Ethics Board (H-01-26-12367) prior to recruitment and data collection. All participants provided informed consent before study participation.

### Study Design

We used a quasi-experimental pre-post within-subject study design to evaluate the WHO TAP module.^20^

### Participants

Participants were recruited using a convenience sampling approach from undergraduate and graduate health sciences programs. Recruitment was conducted using bilingual (English and French) promotional materials distributed through university-approved channels, including faculty recruitment boards, student organization communication platforms, and social media platforms such as LinkedIn and Instagram. Interested individuals followed the study link to the secure, university-hosted REDCap platform, where they completed eligibility screening (Table 1) prior to providing informed consent.

**Table 1.** Inclusion and Exclusion Criteria for Participant Eligibility.

| Inclusion | Exclusion |
| --- | --- |
| Undergraduate and graduate students enrolled in Faculty of Sciences or Faculty of Health Sciences at the University of Ottawa. | Participants who have previously completed the WHO TAP training or similar eHealth capacity-building modules. |
| Aged 18 years or older at the time of consent. | Individuals who are unable to provide informed consent or complete the online module independently. |
| Basic computer literacy and access to a personal device (laptop or tablet) with a reliable internet connection or willingness to use a university-provided device for the duration of the module. |  |
| Interested in learning about APs and mobility aids. |  |

*Note: Participants without access to a personal device were provided with a university computer or tablet to complete the digital module.*

A target sample of 30 participants was established to ensure sufficient power for detecting pre-post changes in self-efficacy, while remaining feasible for the scope of this project. A power analysis was conducted using G*Power 3.1 for a paired-samples t-test comparing two dependent means. With a two-tailed significance level of α = .05, a desired power of .80, and a moderate within-subject standardized mean change of dz = 0.55, the required sample size was estimated at n = 28. Drawing on meta-analytic evidence regarding internet-based instruction in health professions education, the assumed effect size (dz = 0.55) reflected a conservative estimate consistent with medium-sized changes.^21,22^

### Study Protocol: WHO TAP Module

Participants completed the specific WHO TAP module titled “Introduction to Assistive Products,” which provides foundational training on AP provision.^11^ Across five lessons, participants were introduced to core concepts relevant to mobility aid provision, including the role of APs in functioning and participation, the roles of providers and users, and the standardized process of selecting, fitting, using, and following up on APs. As part of the intervention, participants independently progressed through each lesson in sequence, engaged with instructional materials, and completed the embedded interactive components and post-module quiz. Participation occurred entirely online and required approximately two hours.

### Data collection

Before completing the TAP module, participants completed a pre-module self-efficacy questionnaire and a sociodemographic questionnaire collecting age, gender, academic program, year of study, self-identified race/ethnicity, prior experience with APs, and previous exposure to digital learning platforms. Characteristics were selected and adapted based on the PROGRESS-Plus framework, which guides the inclusion of key demographic and social factors that may influence intervention accessibility and outcomes.^23,24^

After completing the TAP module, participants completed three questionnaires.

1. We assessed usability using the System Usability Scale (SUS), a validated 10-item questionnaire assessing perceived usability, including ease of use, efficiency, and satisfaction.^25^ The SUS is often used in digital health research to measure usability and acceptance of the technology.^26^ Items are rated on a 5-point Likert scale and converted to a total score ranging from 0 to 100, with higher scores indicating greater usability.
2. We assessed acceptability using 28 Likert-scale items adapted from the Technology Acceptance Model (TAM) and the Theoretical Framework of Acceptability (TFA). TAM items assessed perceived usefulness (PU), perceived ease of use (PEOU), attitude toward use (ATU), and behavioural intention (BI).^27,28^ TFA items assessed affective attitude (AA), intervention coherence (IC), perceived effectiveness (PEff), burden (B), opportunity cost (OC), and capability (C).^29^ Although the TAM and TFA predate many contemporary eHealth platforms, both frameworks remain applicable and widely used in digital health research.^28,29^ Responses were rated on a 5-point Likert scale, with higher scores indicating greater acceptability.
3. We assessed self-efficacy using an eight-item scale adapted from Bandura’s Multidimensional Scales of Perceived Self-Efficacy (MSPSE), which has been validated across health and educational contexts to measure task-specific confidence.^12,30^ We tailored the items to assess participants’ confidence in performing skills related to mobility aid provision. Participants rated each item on a five-point Likert scale, with higher scores indicating greater perceived self-efficacy.

### Data analysis & interpretation

Analyses were conducted using Jamovi statistical software. Sociodemographic characteristics were analyzed descriptively. Given the modest sample size, subgroup patterns were explored descriptively to contextualize study findings and explore potential equity-related patterns.

Usability scores were calculated according to Brooke’s standardized SUS protocol. For odd-numbered items, 1 was subtracted from the response; for even-numbered items, the response was subtracted from 5. The adjusted scores were summed and multiplied by 2.5 to generate a total score ranging from 0 to 100.^25^ SUS scores were interpreted relative to the established benchmark score of 68, with higher scores indicating greater usability.^31^

Acceptability was analyzed by computing composite mean scores for each TAM and TFA subscale. For each participant, responses to the items within each subscale were averaged to generate one participant-level composite score ranging from 1 to 5. Negatively worded items were reverse coded before averaging so that higher scores consistently reflected greater acceptability. Although individual Likert-scale items are ordinal, composite scores derived from multiple items measuring the same construct were treated as approximately continuous for descriptive analysis.^32^ Unlike the SUS, which requires conversion to a standardized score from 0 to 100, the TAM and TFA subscale scores were retained on the original 1-to-5 scale. Internal consistency reliability was assessed for each subscale using Cronbach’s alpha (α ≥ 0.70 was considered acceptable). Participant-level composite scores were then summarized across the sample using the mean and standard deviation for each subscale. Subscale means were compared descriptively with the neutral scale midpoint of 3, with values above 3 indicating generally more favourable perceptions and values below 3 indicating less favourable perceptions.

For each participant, responses to the eight self-efficacy items were averaged separately at pre- and post-intervention to generate one composite score at each time point. These multi-item composite scores were treated as approximately continuous because they combined responses across related Likert-scale items.^32^ Within-subject change in composite scores was assessed using paired-samples t-tests to determine whether post-intervention scores differed significantly from pre-intervention scores. Statistical significance was evaluated using a two-tailed alpha of 0.05, with exact p-values reported. Normality of the difference scores was assessed using Shapiro-Wilk tests; when assumptions were met, parametric testing was retained. The magnitude of change was quantified using Cohen’s dz effect size, and 95% confidence intervals were reported for both mean differences and effect sizes. In addition to composite analysis, item-level changes across competencies were examined descriptively.

## Results

### Sociodemographic characteristics

A total of 53 participants initiated the study through the REDCap platform. Of these, 33 participants were included in the final analysis after completing both pre- and post-intervention questionnaires, corresponding to a completion rate of 62%. The sample consisted primarily of undergraduate students (n = 20, 60.6%) predominantly in upper years of study (years 3-4; n = 17, 51.5%) and graduate-level participants (n = 13, 39.4%). The sample was largely composed of women (66.7%), participants aged 18-24 years (93.9%), and students in health-related disciplines (Table 2).

**Table 2.** Sociodemographic Characteristics of Study Participants (N = 33)

| Characteristics | Category | n (%) |
| --- | --- | --- |
| Gender | Woman | 22 (66.7) |
|  | Man | 9 (27.3) |
|  | Prefer not to say | 2 (6.1) |
| Age | 18–24 years | 31 (93.9) |
|  | 25–34 years | 2 (6.1) |
| Program of Study | Health Science | 16 (48.5) |
|  | Biomedical Sciences | 8 (24.2) |
|  | Rehabilitation Sciences | 9 (27.3) |
| Year of Study | Undergraduate Year 2 | 3 (9.1) |
|  | Undergraduate Year 3 | 7 (21.2) |
|  | Undergraduate Year 4 | 10 (30.3) |
|  | Graduate (Master's) | 12 (36.4) |
|  | Graduate (PhD) | 1 (3.0) |
| Ethnicity | Canadian | 14 (42.4) |
|  | South Asian | 10 (30.3) |
|  | East Asian | 2 (6.1) |
|  | Middle Eastern | 3 (9.1) |
|  | African | 2 (6.1) |
|  | Prefer not to say | 2 (6.1) |
| **Language** | English | 28 (84.8) |
|  | French | 5 (15.2) |

### Usability of the e-Health Training Tool

Among the 33 participants, the mean score on the SUS was 76.97 (SD = 9.66; range = 52.50-97.50), exceeding the established benchmark of 68 and indicating above-average usability, with scores approaching the threshold for excellent usability. The distribution of individual SUS scores (Figure 1) showed that most participants scored above the benchmark, with scores largely clustered between 70 and 90. A small number of participants (n = 4) had lower scores (< 68), which contributed to some variability but represented a minority of responses.

**Figure 1.**
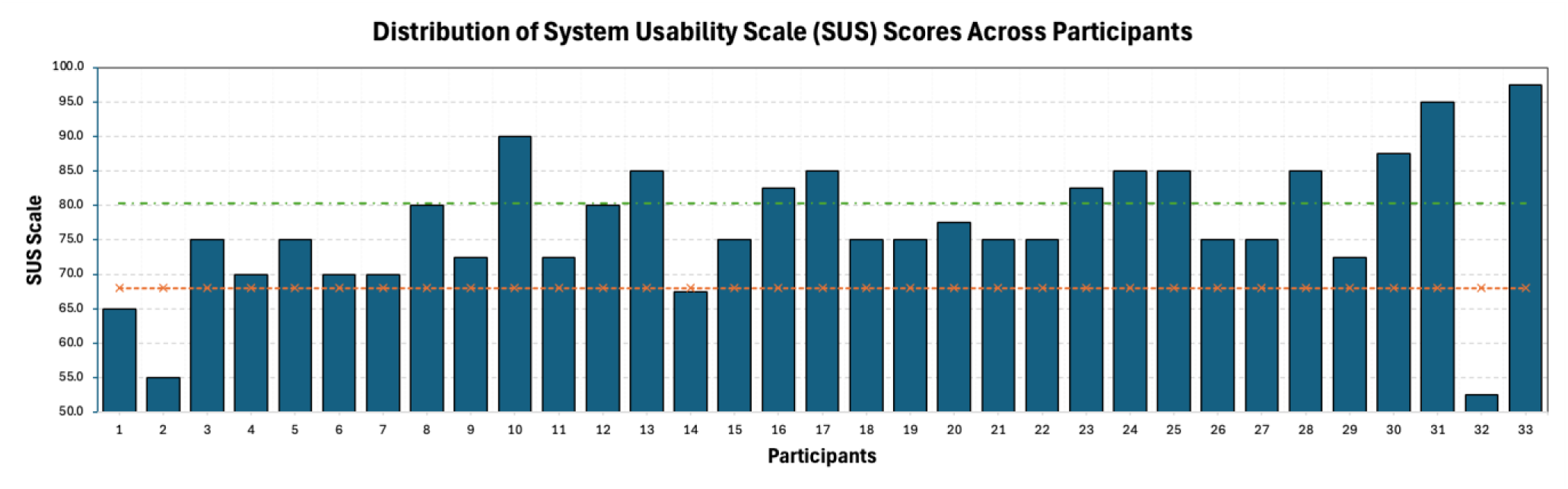
Distribution of SUS scores across participants (N = 33). Each bar represents an individual participant’s SUS score (0–100). The dashed orange line indicates the average usability benchmark (68), and the dashed green line indicates the threshold for excellent usability (80.3), based on established SUS interpretation guidelines.

### Acceptability of the e-Health training tool

Overall acceptability was high (M = 3.9), with all subscales exceeding the neutral midpoint of 3 on a 5-point Likert scale. Aggregate scores were similar for the TAM (M = 3.9) and TFA (M = 3.9) subscales (Figure 2). Within TAM, perceived ease of use (M = 4.2) and perceived usefulness (M = 4.0) showed the highest scores, while attitude toward use (M = 3.6) and behavioural intention (M = 3.6) were slightly lower but remained above the midpoint. Within TFA, intervention coherence (M = 4.6) demonstrated the highest score, followed by perceived effectiveness (M = 4.2) and capability (M = 4.3). Affective attitude was moderate (M = 3.7). Burden (M = 3.3) and opportunity cost (M = 3.3) had comparatively lower scores, though still above the midpoint.

**Figure 2.**
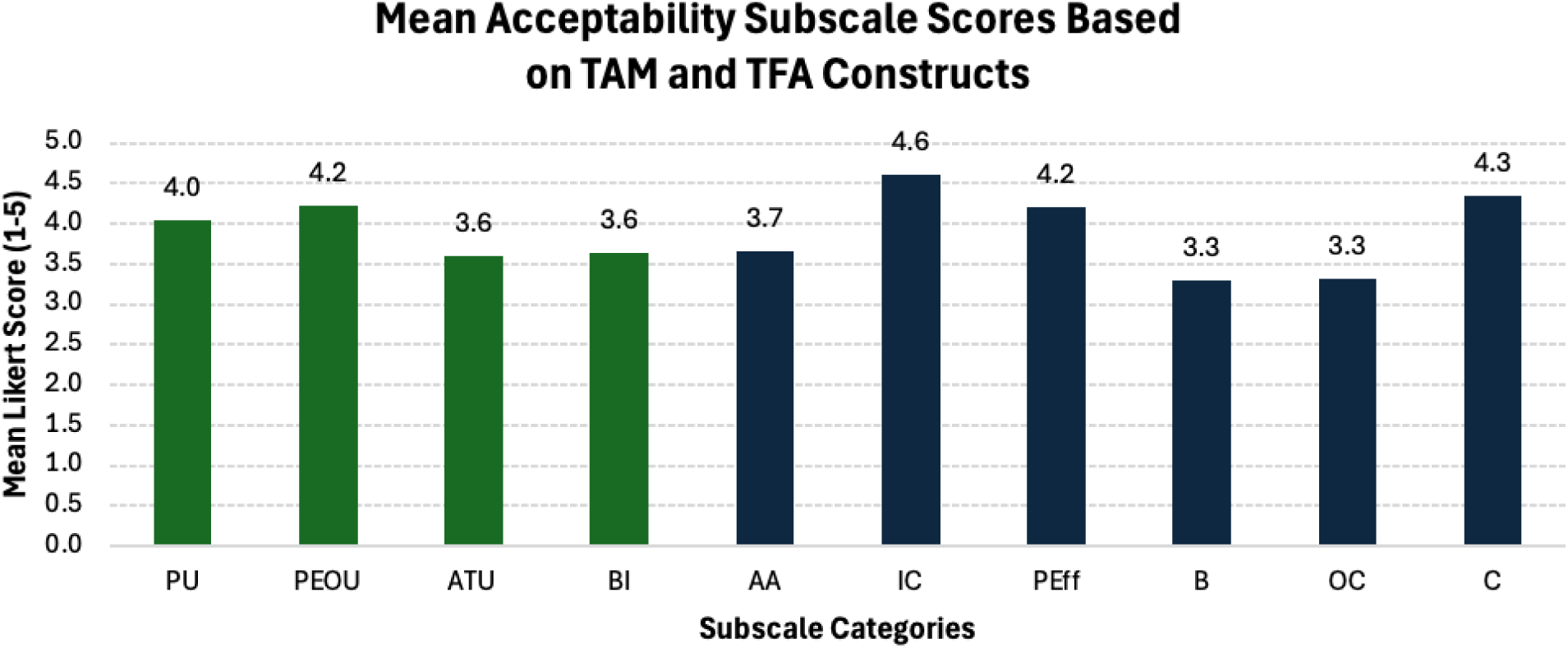
Acceptability of the intervention based on TAM (green) and TFA (dark blue) constructs. PU, Perceived Usefulness; PEOU, Perceived Ease of Use; ATU, Attitude Toward Use; BI, Behavioural Intention; AA, Affective Attitude; IC, Intervention Coherence; PEff, Perceived Effectiveness; B, Burden; OC, Opportunity Cost; C, Capability.

### Changes in Self-efficacy

Mean self-efficacy scores increased across all eight mobility aid-related competencies following completion of the WHO TAP module (Table 3). Pre-training mean scores ranged from 1.55 to 3.39, while post-training scores ranged from 3.82 to 4.42. All items demonstrated increases in mean scores, with the largest absolute changes observed in Q7 (Δ = 2.82), Q6 (Δ = 2.39), and Q3 (Δ = 2.21). At the aggregate level, mean self-efficacy scores increased from 2.32 ± 0.58 pre-intervention to 4.23 ± 0.36 post-intervention (Δ = 1.90).

**Table 3.**
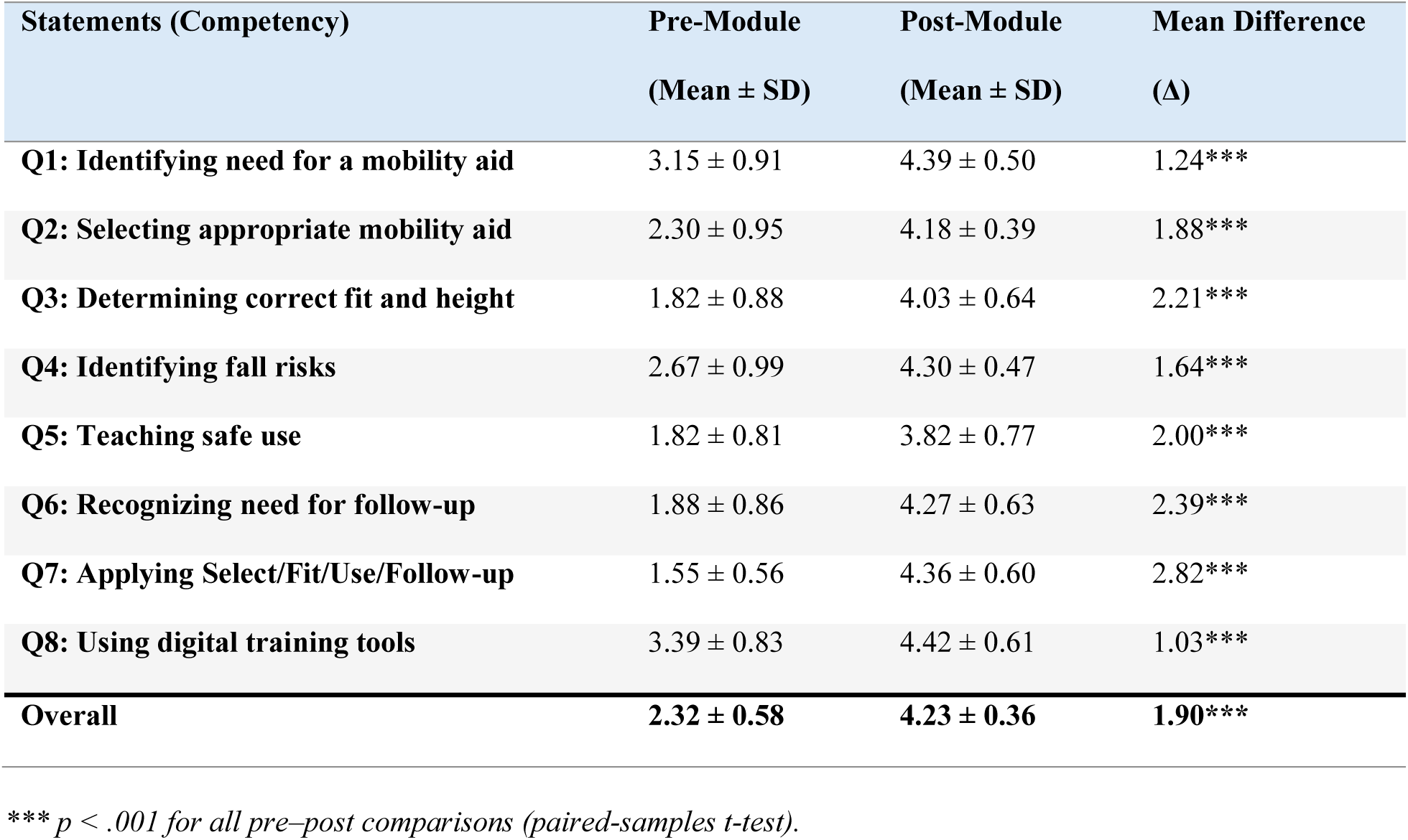
Changes in Self-Efficacy Scores Pre- and Post-Training (Mean ± SD)

| Statements (Competency) | Pre-Module<br>(Mean ± SD) | Post-Module<br>(Mean ± SD) | Mean Difference<br>($\Delta$) |
| --- | --- | --- | --- |
| **Q1: Identifying need for a mobility aid** | 3.15 ± 0.91 | 4.39 ± 0.50 | 1.24*** |
| **Q2: Selecting appropriate mobility aid** | 2.30 ± 0.95 | 4.18 ± 0.39 | 1.88*** |
| **Q3: Determining correct fit and height** | 1.82 ± 0.88 | 4.03 ± 0.64 | 2.21*** |
| **Q4: Identifying fall risks** | 2.67 ± 0.99 | 4.30 ± 0.47 | 1.64*** |
| **Q5: Teaching safe use** | 1.82 ± 0.81 | 3.82 ± 0.77 | 2.00*** |
| **Q6: Recognizing need for follow-up** | 1.88 ± 0.86 | 4.27 ± 0.63 | 2.39*** |
| **Q7: Applying Select/Fit/Use/Follow-up** | 1.55 ± 0.56 | 4.36 ± 0.60 | 2.82*** |
| **Q8: Using digital training tools** | 3.39 ± 0.83 | 4.42 ± 0.61 | 1.03*** |
| **Overall** | **2.32 ± 0.58** | **4.23 ± 0.36** | **1.90\*\*\*** |

\*\*\* $p < .001$ for all pre-post comparisons (paired-samples $t$ -test).

A paired-samples t-test indicated that post-intervention self-efficacy scores were significantly higher than pre-intervention scores, t(32) = −16.00, p < .001. The magnitude of change was large (Cohen’s dz = −2.79, 95% CI [−3.55, −2.03]), indicating a substantial increase in perceived self-efficacy following training. The assumption of normality was met (Shapiro-Wilk W = 0.957, p = .218) (Figure 3a-b).

**Figure 3a.**
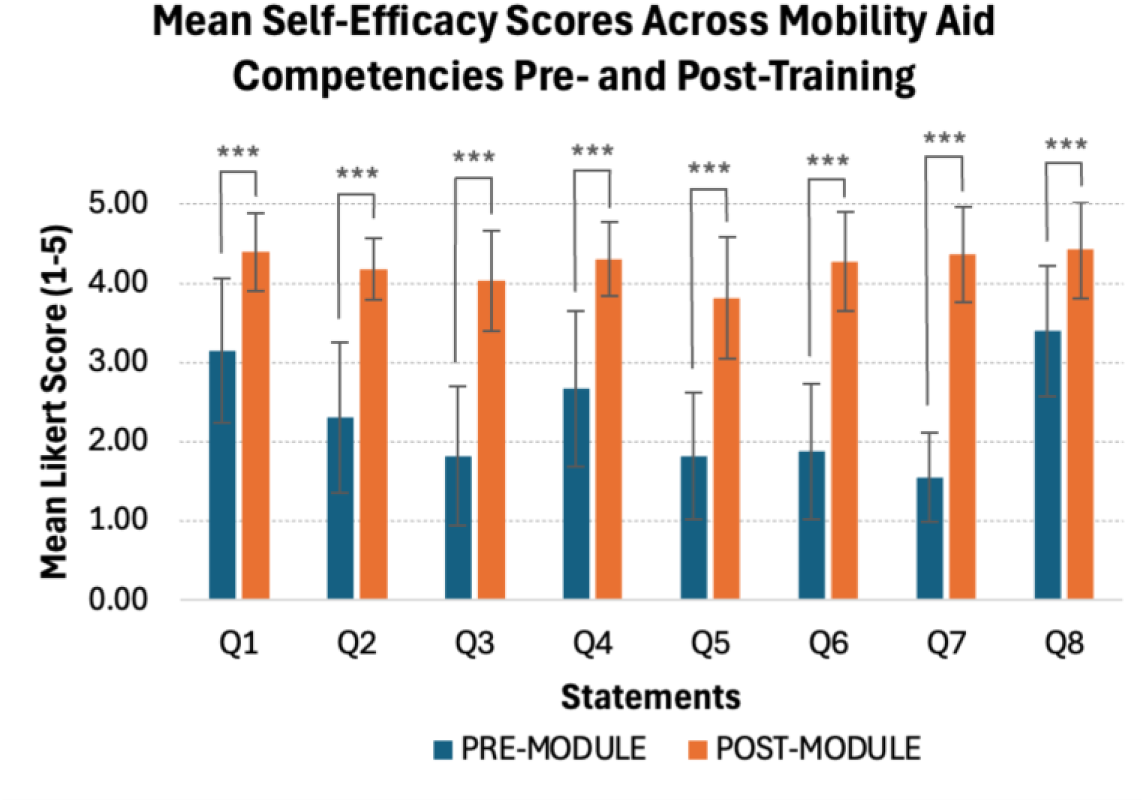
Changes in self-efficacy following training. Error bars represent standard deviation. Q1-Q8 correspond to mobility aid provision competencies. ***p < .001, indicating statistically significant pre-post differences.

**Figure 3b.**
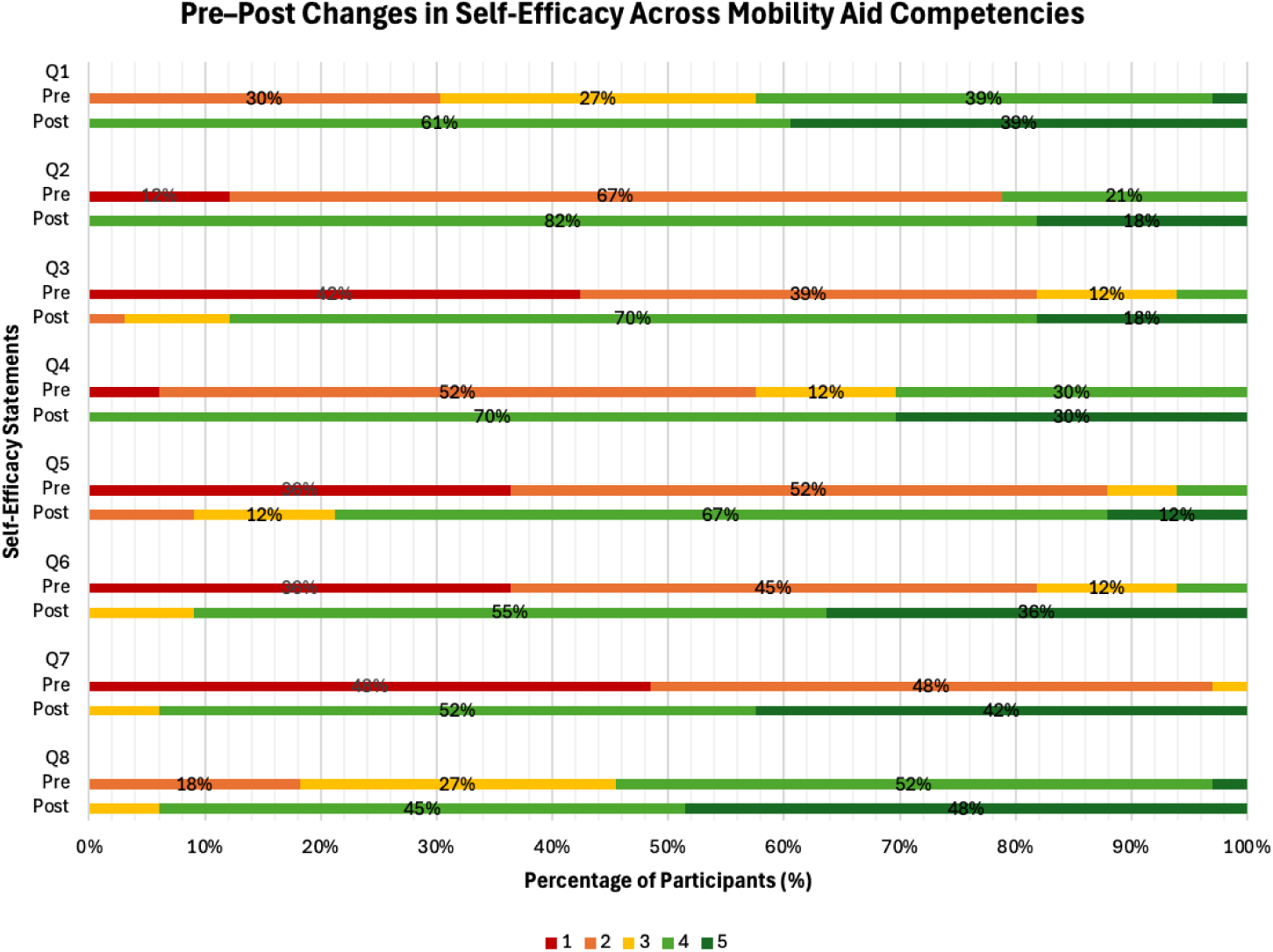
Pre-post changes in self-efficacy across mobility aid competencies. Distribution of Likert-scale responses (1-5) for each competency (Q1-Q8) before and after training, showing a shift toward higher agreement categories post-training.

Changes in response distributions demonstrated that pre-training responses were primarily concentrated in lower Likert categories (1-2), with a shift in post-training responses toward higher agreement categories (4-5) across all competencies (Figure 3b).

## Discussion

Training personnel to provide assistive products safely and effectively is essential to improving access, and eHealth training may help extend foundational education to a broader workforce. Results from this study suggest that the WHO eHealth TAP training module was perceived as usable and acceptable and was associated with increased self-efficacy among participants. WHO guidance on digital interventions for health system strengthening frames digital learning as a complement to conventional training and, consistent with research on healthcare workers’ workload, highlights the need to assess intervention feasibility and acceptability, ensure equitable access, and provide supportive implementation conditions, including infrastructure, technical support, workflow integration, staff readiness, and organizational commitment.^33,34^ From an implementation perspective, these findings support eHealth modules as a promising approach to standardizing foundational workforce training, particularly when integrated into existing health system structures to support long-term sustainability.

### Usability of the e-Health training

The mean SUS score exceeded the established benchmark, suggesting that participants navigated the module with relatively little friction and that its usable interface may have reduced unnecessary mental effort related to navigation, allowing learners to focus more directly on the training content.^35^ Most participants scored above the benchmark, suggesting that the module was generally perceived as intuitive, learnable, and easy to navigate. Taken together, these findings suggest that the interface supported learning for most participants, while minor usability barriers may have remained.

### Acceptability of the e-Health Training

The acceptability of the WHO eHealth TAP training module reflects a coherent “high-value, low-friction” pattern, whereby participants perceived the training as both useful and easy to use, while also demonstrating a clear understanding of its purpose and feeling capable of applying its content. Within the TAM, high ratings for perceived usefulness and perceived ease of use are positive indicators of future technology adoption and strongly influence downstream outcomes such as attitude and behavioural intention to use.^36,37^ However, the comparatively lower ratings for attitude toward use and behavioural intention suggest that even strong initial positive perceptions of usefulness and ease of use may not always directly translate into stronger intentions to continue using the module. This pattern may reflect external or contextual constraints, such as time demands, competing priorities, or workflow-related factors, which are not fully captured by core TAM constructs but are recognized in broader models of technology adoption.

From a TFA perspective, the strong endorsement of intervention coherence, perceived effectiveness, and capability indicates that participants understood the intervention, believed in its value, and felt confident in their ability to engage with and apply its content.^29^ In contrast, the comparatively lower scores for burden and opportunity cost suggest that practical considerations, such as the time and effort required to complete the module, may have been viewed less positively.

### Self-efficacy of the e-Health Training

Pre-post improvements in self-efficacy across all mobility aid competencies suggest that the module supported participants to feel able to apply what was learned. Within social cognitive theory, self-efficacy is not a trivial perception; it influences whether individuals initiate behaviour, the effort they invest, and their persistence when faced with challenges.^38^ This distinction is especially relevant in the context of assistive device provision, where efficacy in tasks such as selecting, fitting, training, and follow-up may act as an important precursor to future clinical applications.

These findings are consistent with prior literature in wheelchair and mobility training, which shows that targeted educational interventions can improve learner confidence, skill capacity, and perceived readiness to apply training in practice, including in remote and hybrid formats.^39,40^ Broader evidence also suggests that while training approaches vary widely, integration into professional education remains inconsistent across settings, reinforcing the importance of scalable and accessible training models such as eHealth modules.^41^ A plausible interpretation is that the high usability and acceptability observed in this study may have contributed to self-efficacy gains by allowing participants to focus on learning rather than navigating the platform. This should be interpreted cautiously, as the present study design does not support causal conclusions. However, this interpretation aligns with existing theory suggesting that when learners perceive a training tool as understandable, useful, and coherent, they are more likely to engage with the material at a depth sufficient to strengthen confidence.^42^

Increased self-efficacy should not be equated with demonstrated competence. While the present findings indicate improved perceived preparedness, conclusions regarding actual clinical proficiency require further investigation using objective performance-based measures and an evaluation of effects on patient outcomes. Nevertheless, the consistent improvements observed across all competency domains suggest broad skill development rather than isolated gains, supporting the potential of the module to enhance learner readiness in AP provision.

### Strengths & Limitations

This study has several strengths. First, the use of a multi-framework evaluation approach, integrating the KTA framework with usability (SUS), acceptability (TAM and TFA), and self-efficacy (MSPSE), allowed for a comprehensive and theory-informed assessment of the intervention and future implementation. This evaluation approach provides a holistic understanding of how participants interacted with and responded to the training. It also contributes to the early phases of the action cycle by assessing key factors that influence how an intervention can be effectively adapted to different contexts. Finally, the focus on competency-based outcomes strengthens the relevance of the findings, as improvements were observed across multiple domains rather than isolated skill areas.

Several limitations should also be considered. First, the reliance on self-reported measures introduces the possibility of response bias, including social desirability or immediate post-training enthusiasm.^43^ To minimize this, data were collected anonymously using standardized and validated instruments, which may reduce pressure to respond in socially desirable ways and support more consistent reporting across participants.^44^ Second, the lack of follow-up monitoring prevents conclusions on durability of learning, sustained engagement with the platform, or translation into actual clinical behaviour.^45^ From a KTA perspective, this reflects the early-stage nature of the study, as later phases of the action cycle would require evaluation of longer-term use and real-world outcomes. Third, participants were undergraduate and graduate students rather than the community- and primary-level healthcare workers for whom TAP was primarily developed. Although this student sample was appropriate for an early-stage evaluation of the module, the findings may not generalize to TAP’s intended users, particularly personnel working in low-resource or community-based settings. Future research should evaluate the module among its intended users and within real-world service-delivery contexts. Finally, while the MSPSE was adapted to reflect mobility aid competencies, further validation within this specific context would strengthen the interpretability of the self-efficacy findings.

### Future Directions

Future research should first extend evaluation beyond immediate learner outcomes to examine longer-term impacts on clinical behaviour, patient outcomes, and service delivery. Studies should examine whether individuals trained through the module are able to apply AP provision steps effectively in practice, communicate clearly with users, support safe use, and provide follow-up.

Evaluating AP user satisfaction, device fit, functional outcomes, and rates of device abandonment would help determine whether eHealth training ultimately improves the quality of AP provision. This highlights the importance of moving from early-stage evaluation toward real-world implementation and impact assessment. These considerations are especially important in low-resource contexts, where infrastructure and workflow constraints may differ substantially from university-based environments.^3,41^ Overall, the findings reinforce the potential of digital training tools as part of broader system-level strategies to strengthen workforce capacity, provided that they are implemented in ways that are contextually relevant, sustainable, and aligned with clinical practice.

Accordingly, future implementation efforts should consider contextual factors, particularly when extending beyond a student sample to community health workers operating in diverse socioeconomic settings. These factors include protected learning time, offline or low-bandwidth functionality, local language adaptation, and alignment with existing service-delivery pathways.

## Conclusion

This study provides an early-stage evaluation of the WHO eHealth Training in Assistive Products (TAP) module, demonstrating high usability, favourable acceptability, and significant improvements in self-efficacy across mobility aid competencies. Together, these findings suggest that the module not only supports positive user experiences but also strengthens learners’ self-efficacy to apply key skills in practice. Importantly, the integration of usability, acceptability, and self-efficacy highlights the value of a multi-dimensional evaluation approach, showing that effective digital training extends beyond content delivery to include how learners interact with, perceive, and apply the material. These findings reinforce the potential of eHealth training tools to support workforce capacity-building, particularly in contexts where access to traditional training is limited. While further research is needed to examine long-term impacts on clinical practice and patient outcomes, this study provides foundational evidence supporting the role of digital training in strengthening AP provision. Overall, the findings highlight the promise of scalable, context-sensitive eHealth interventions as part of broader strategies to advance global health equity and improve access to rehabilitation services.

## Acknowledgements

The authors have no acknowledgements to declare. AI tools were used to support writing clarity and grammar editing. The authors independently reviewed and approved all content and take responsibility for the manuscript’s ideas, analyses, and interpretations.

## Data Availability

Data generated and analyzed during this study are not publicly available due to research ethics considerations but may be available from the corresponding author upon reasonable request.

## Contributions

The first author developed and conducted the study methodology with guidance from Dr. O’Neil. The first author led the data analysis, interpretation of results, and manuscript preparation, which Dr. O’Neil and Dr. Sauvé-Schenk reviewed.

